# A Needs Analysis of Parents Following Sudden Cardiac Death in the Young

**DOI:** 10.1101/19000877

**Authors:** Kristie McDonald, Louise Sharpe, Laura Yeates, Christopher Semsarian, Jodie Ingles

**Affiliations:** Agnes Ginges Centre for Molecular Cardiology at Centenary Institute, University of Sydney, Australia; School of Psychology, University of Sydney, Sydney Australia; Sydney Medical School, University of Sydney, Sydney, Australia; Department of Cardiology, Royal Prince Alfred Hospital, Sydney, Australia

**Keywords:** Sudden cardiac death, needs analysis, psychological wellbeing

## Abstract

The sudden cardiac death (SCD) of a young person is a devastating event for any parent. Inherited heart disease is often either identified or assumed to be the cause. Few studies have explored the psychosocial impact to the surviving at-risk family members. We sought to investigate the needs of parents who have experienced the SCD of their child (≤45 years). A quantitative needs analysis questionnaire was developed based on semi-structured interviews, including one focus group, and a review of relevant literature. There were 38 parents who completed a cross-sectional quantitative survey. Parents’ perceived needs for information and support spanned medical, psychosocial, spiritual and financial domains. Medical information and support needs were identified as the most important domains, followed by psychosocial, spiritual and financial information and support needs. Importantly, psychosocial information and support needs were reported as the most unmet need, endorsed by 54% of parents. Medical information and support needs were reported as unmet by almost one third of parents. The two most endorsed needs were “To have the option of whether or not you would pursue genetic testing for yourself or family members” and “To understand what happened”. This work demonstrates for the first time, the multifactorial needs of parents after SCD in the young. With the greatest unmet need reported as psychosocial needs, there is clear necessity to find ways of integrating psychological support in to the care of families after SCD in the young.

**KEY QUESTIONS:** *What is already known about this subject?:* There is currently very little known about the needs of families following a sudden cardiac death due to inherited heart diseases. We know there is significant risk of poor psychological outcomes including posttraumatic stress and prolonged grief, even years after the death, however this is one of the first studies to formally evaluate the needs of parents during this time.

*What does this study add?:* We show that medical information and support needs are ranked very highly, but psychosocial support needs are the most unmet. Our findings provide a platform for developing an approach to delivering psychosocial support interventions in this population.

*How might this impact on clinical practice?:* Currently clinical and research efforts in this setting focus on clinical and genetic aspects of care. Here we show the critical need to also focus on the psychological care needs in this population. These data will help to guide services in integrating psychological support in to their multidisciplinary clinic models.

## INTRODUCTION

The loss of a young family member is one of the most tragic events for an individual to endure, particularly when the death is sudden, unexpected, unexplained and possibly heritable. Sudden cardiac death (SCD) in the young has an annual incidence of 1.3 per 100,000 individuals in Australia and New Zealand every year.^1^ Inherited heart diseases account for a large majority of SCD in the young, with 40% being unascertained at postmortem examination and presumed due to inherited arrhythmia syndromes, while identifiable structural causes such as hypertrophic cardiomyopathy, arrhythmogenic right ventricular cardiomyopathy and dilated cardiomyopathy make up approximately 16% of cases.^1^

First-degree relatives who have experienced the SCD of a young relative report significantly greater stress, anxiety and depression compared to the general population.^2^ Clinically significant symptoms of posttraumatic stress and/or prolonged grief were reported by almost half of family members, on average 6 years after the death.^3^ Whilst these findings provide a brief insight into the psychological difficulties experienced by families, we are yet to understand a family’s needs in the wake of the SCD of a young person.

Prior research to understand the needs of Swedish parents after the SCD of their 15-35 year old child identified four needs using a qualitative approach. These included evidence of the death, understanding the circumstances through reconstruction, medical explanation, and a sensitive approach by professionals.^4^ The focal goal of being understood by health professionals and themselves understanding what was going on was highlighted as important. With greater emphasis and awareness of the familial nature of SCD in the young, we sought to comprehensively understand the needs of parents whose child (≤ 45 years) experienced a SCD that was thought to be due to an inherited heart disease, through development and distribution of a needs questionnaire.

## METHODS

### Participants

This was a cross-sectional survey study. Parents were recruited from the Genetic Heart Disease Clinic, Royal Prince Alfred Hospital (RPA) in Sydney, Australia. We included parents whose child had died suddenly aged ≤ 45 years with an inherited heart disease identified (or presumed to be the cause) at postmortem examination. Only cases where there was no premorbid diagnosis of an inherited heart disease were eligible. We required a sufficient level of written English skills, as self-nominated by the participant. The local human research ethics committee approved this study and all participants gave written informed consent prior to commencement of the interviews and questionnaire.

### Questionnaire Development

We developed the *SCD Parental Needs Questionnaire* based on one semi-structured focus group and nine semi-structured interviews. Thematic analysis was employed to elicit descriptive data concerning the needs that parents had experienced from their own perspective. The needs identified by parents who were interviewed and other previously designed needs assessments including in oncology,^5^ cardiac patients^6^ and parents bereaved in pediatric intensive care literature^7^ were reviewed to develop the content and format of the needs assessment. The questionnaire included an inventory of 45 items assessing the met and unmet needs reported by parents after SCD in the young. Forty-four needs questions were presented to each respondent. A single screening question was presented to each respondent, assessing whether a gene variant was identified, and for parents whom answered ‘yes’ to this, an additional five follow up needs questions were displayed. Each needs question had a dichotomous yes/no response. If ‘yes’ was selected, an additional two items were presented, assessing the importance of the need, measured on a three-point scale (low, moderate, high) and whether the need had been met (Figure 1). One of the 45 items was a screening question and only relevant to parents for whom a gene variant was identified. For this item, if parents selected ‘yes’ from the dichotomous response, an additional five follow up questions were shown, in the same format as all other need questions.

**FIGURE 1:**
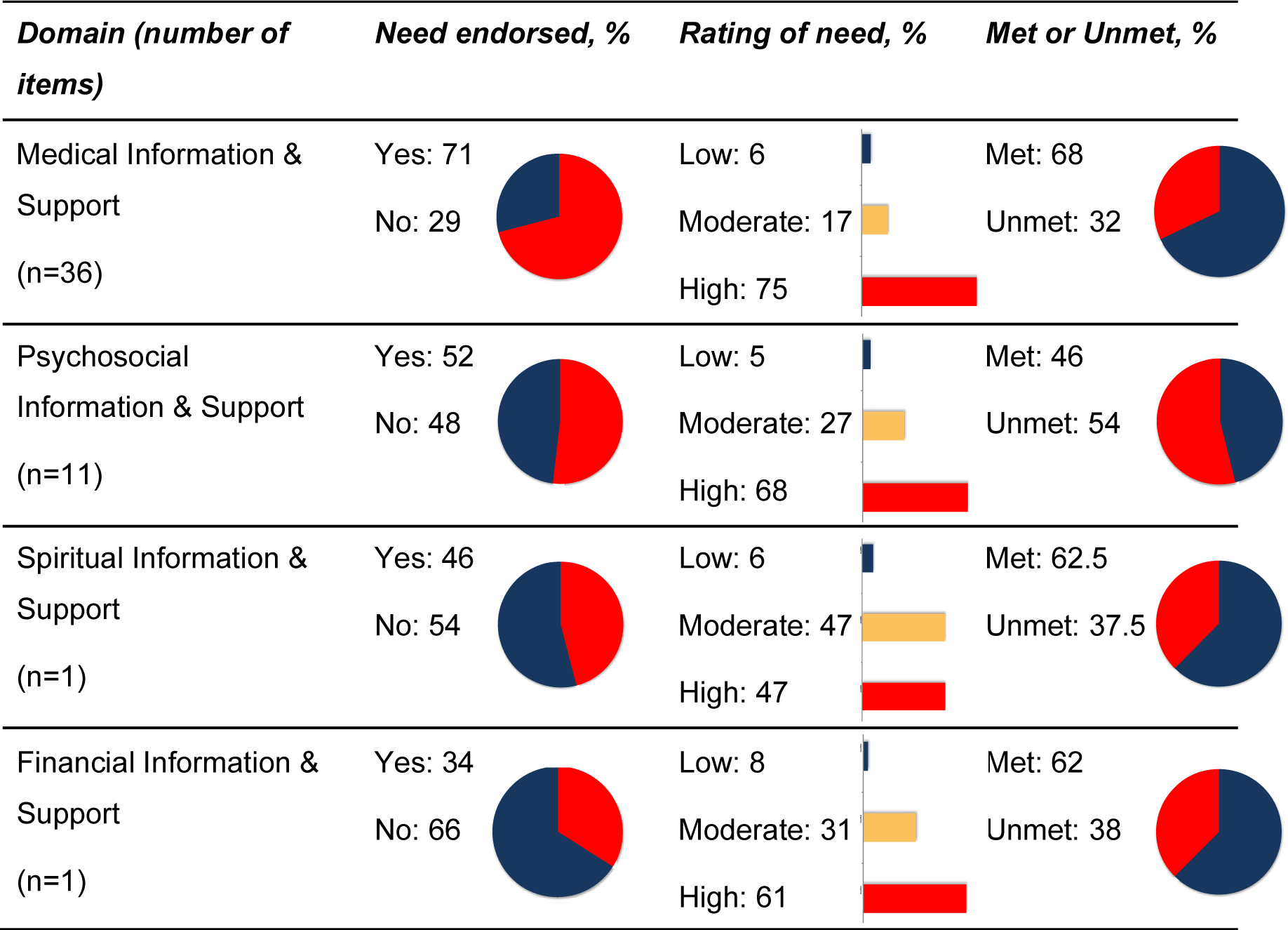
Summary of needs by domain.

In addition, demographic questions were included about the respondent (name, age and marital status), and their deceased child (gender, age at death, year of death and cause of death). Statistical analyses were conducted using *SPSS v21*. Missing data were not included. Descriptive statistics were used to investigate frequency of endorsed need, the importance and whether the needs were met.

### SCD Parental Needs Questionnaire

Based on the qualitative analyses, 4 domains of need were included in the *SCD Parental Needs Questionnaire*:

1. Medical information and support needs
2. Psychosocial information and support needs
3. Financial information and support needs
4. Spiritual information and support needs

#### 1. Medical information and support needs

This domain encompassed parent’s interactions with medical professionals at the time of and in the aftermath of the death. Professionals included paramedics, police, medical doctors, nurses and administrative support, and those involved in later care including general practitioners and cardiac specialists. Six sub-categories were established: 1) information about cause of death; 2) information about heritability and consequent discussion of genetic testing; 3) information about the medical ramifications for self and family; 4) appropriate and timely referrals e.g., to cardiac specialists and/or genetic counsellors; 5) opportunity to view the body and 6) an empathic and supportive approach by all.

#### 2. Psychosocial information and support needs

This domain described psychological and social support identified by parents. Within this domain, six sub-categories were established; 1) support from family; 2) support from friends; 3) support from work, including time off work and understanding of ongoing impact; 4) support from the community, including understanding by school of impact on siblings; 5) information about and access to individual therapy and 6) information about support groups.

#### 3. Financial information and support needs

This domain was utilized to comprise monetary needs. Two sub-categories were established: 1) needs relating to the young person’s estate (both dealings with insurance and bank accounts) and 2) medical costs (including clear emphasis on what appointments and tests were reimbursed by insurance).

#### 4. Spiritual information and support needs

This domain encompassed all needs participants identified as relevant to: 1) their own religiosity/spirituality; 2) their family’s religiosity/spirituality and 3) their child’s religiosity/spirituality.

## RESULTS

### Participants

One hundred and twenty-four parents were directly approached, with social media recruitment used to increase participation. A total of 38 parents completed the questionnaire. Participants were approached through postal mail (19% response rate; 24/124), telephone (12% response rate; 3/25), email (42% response rate; 11/26) and a Facebook advertisement (n = 2). Characteristics of the participants and the decedent are shown in Table 1. There were 8 (21%) male participants and mean age was 59 ± 11 years. In terms of the decedent, the mean age at death was 23 ± 10 years and 28 (74%) were male. The time since death ranged from 2-17 years and 26 (68%) had no clear cause of death identified at postmortem examination.

**TABLE 1:**
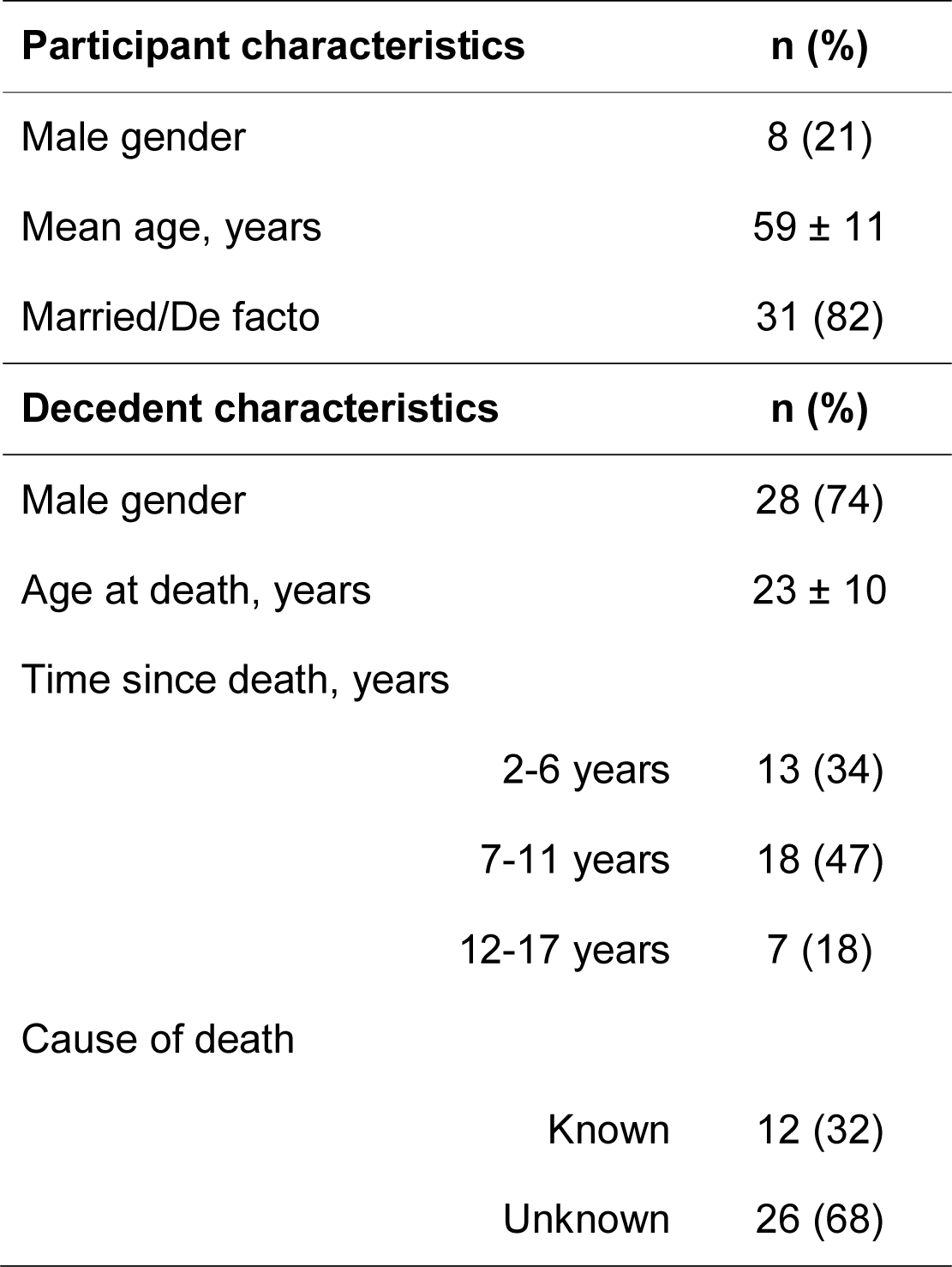
Characteristics of the parents and decedent.

### Needs across the domains

The mean of the responses across the 4 domains are shown in Figure 1. Of the 36 items relating to medical information and support needs, on average 71% endorsed the items as a need and 75% rated these needs as having high importance. Overall 32% reported at least one need in the medical information and support domain as unmet. There were 11 items relating to psychosocial information and support, with on average 52% reporting these as a need and of those, 68% felt the importance was high and 54% report these needs as unmet. The spiritual need was ranked as the 38^th^ most important need, being endorsed by 46% of parents, and 94% subsequently rating it as of moderate to high importance. There were 38% of parents who reported this need as unmet. The financial need was rated as the 46^th^ most important need, being endorsed by 34% of parents; with 62% rating the importance as high and 38% reported this need as being unmet.

### Frequently endorsed needs

All needs items are shown in Supplementary Table 1. Twelve needs were endorsed by ≥80%. Of these, 11 were medical information and support needs, while one was a psychosocial information and support need. Between 63%-93% reported these 12 needs as being of “high” importance. The two most endorsed needs were “To have the option of whether or not you would pursue genetic testing for yourself or family members”, with 33% identifying this need as unmet; and “To understand what happened”, which was reported unmet by 60% of participants.

### Unmet needs

Psychosocial needs were considered the most unmet. The items “For support from your family” and “For access to professional counselling or psychological services to deal with your bereavement” were reported as needs by 89% and 74%. The most unmet needs were “For a health professional or resources to provide you with advice for dealing with people who expected you to be ‘back to normal’ after your child’s death” by 75%; and “For medical professionals to provide you with the opportunity to ask questions about your child’s death” and “For referral to someone to talk to who understands and has been through a similar experience” by 69%. Of note, the item “To understand the genetic cause of your child’s death” was regarded as a need by 72%, and 67% of participants reported this as unmet.

## DISCUSSION

We show the greatest needs of families after SCD in the young are medical information and support needs, particularly relating to understanding the cause of the death. The greatest unmet needs were focused in the psychological domain, with participants reporting a need for health professionals, family and friends to have a clearer understanding of the unique clinical experience they are confronted with. As we better elucidate the clinical and genetic underpinnings of SCD in the young, our study suggests optimising the specialised multidisciplinary clinic model could potentially have an important impact on ensuring needs of surviving family members are met.

All needs in our survey were endorsed by at least 26% of participants, indicating their overall relative importance. Medical information and support needs were the most commonly endorsed overall, consistent with previous research highlighting that sensitive and empathic communication of the reason for the death is of paramount importance for parents.^4, 8^ Understanding the cause of death was considered a key need for many, and overall was largely unmet. Given the unexpected nature of SCD in the young, whereby a large proportion of deaths remain unexplained, this is not surprising but puts even greater emphasis on the positive impact that our continuing research efforts to understand the causes of SCD in the young will have. In particular, many participants indicated access to genetic testing or understanding the genetic cause to be an important need, highlighting the importance of maintaining realistic expectations regarding the diagnostic yield of postmortem genetic testing with our families.^9^ At present, this is likely not greater than 15% and there is a high likelihood of uncertain genetic findings especially with increasing gene panel sizes.^1, 10^

While there was a clear need for understanding the cause of death, it should be noted that these families were recruited from a specialised multidisciplinary clinic where there is a strong interest in understanding SCD in the young. It could be argued this patient group have had even greater opportunity for a cause of death to be established, access to experienced health professionals including cardiac genetic counsellors who develop a relationship with the family and provide in-depth pre- and post-test genetic counselling. The needs relating to understanding the cause of death, including genetic testing options are therefore likely underestimated in our study.

Management of families after a young SCD ideally should be performed in a specialised multidisciplinary clinic, including cardiologists and genetic counsellors to explore clinical screening and genetic testing options. There are known challenges commonly encountered when caring for families after SCD of a young relative. In particular there is a high prevalence of clinically significant symptoms of posttraumatic stress and prolonged grief.^3^ Reluctance to undergo clinical surveillance is also not uncommon, especially amongst mothers of the decedent.^11, 12^ Cardiac genetic counsellors have evolved to play a key role in the management of families after SCD in the young.^13^ Indeed, previous research has highlighted the benefits of genetic counselling, conceptualising it as a highly circumscribed form of psychotherapy.^14^ Thus, increased emphasis on genetic counselling is essential to continuation of care and communication with health services. Of note, one critical role for genetic counsellors in this setting should be identification of individuals who require assessment by a clinical psychologist.

Psychosocial needs were, on average, the domain of second most importance and overall considered the most unmet. Such results shed light on the current gap in service provision, with the implication that parents are asking for, and not receiving, sufficient psychosocial support to help cope with bereavement. Whilst previous research has both proposed the need for and demonstrated the benefits of a multidisciplinary team in the care of families with inherited cardiovascular diseases,^12^ further research is needed to understand how to best deliver psychosocial support to SCD parents and their families. Exploring approaches such as individual counselling, for example meaning-making narrative therapy,^15^ and the benefits of support groups such as affiliation and emotion support through a sense of belonging, instrumental support in the form of a safe place for discussion, practical aid in the form of informational support and normalisation and social comparison, may be beneficial.^16^ The role of family and friends in providing support to grieving individuals is already well-described.^17^ Examining ways this could be further encouraged or explored with families may likewise be helpful.

When considering how to communicate information effectively to parents, educational interventions such as the GRIEV_ING death notification protocol^18^ may be useful within the SCD context. The GRIEV_ING mnemonic encourages professionals to ‘*G*ather’ all family members, provide ‘*R*esources’, ‘*I*dentify’ yourself, your patient and the state of knowledge, ‘*E*ducate’ the family about the events, ‘*V*erify’ that death has occurred, *‘_’* provide space to process and absorb information, ‘*I*nquire’ about questions, provide ‘*N*uts and bolts’ about practical tasks (e.g., organ donation and funeral services), and ‘*G*ive’ access to information and support for questions that may arise later. Given the limited understanding of the cause of death, there may be limited ability to confidently and empathically provide information and support. Future research may evaluate how such an educational intervention model can be utilized during later stages of grieving, when cause of death is established or remains unascertained. Additionally, future research may benefit from exploring how families cope with the ambiguity surrounding SCD.

Our study was limited by the modest sample size, and despite best efforts the response rate was relatively low. The difficulty and sensitivities of recruiting this unique patient population cannot be underestimated, and the final results provide a framework for considering what needs are considered important. How these might apply more broadly, particularly to the population who do not have access to specialised clinics should be examined.

## Conclusion

Understanding the multifactorial needs of parents after SCD in the young is paramount for informing health care professionals in how to best assist and support this vulnerable population. Unmet psychosocial needs shed light on the current gap in service provision, with the implication that parents are asking for, and not receiving, sufficient psychosocial support to help cope with bereavement. Identifying how to best deliver psychosocial support to families is important, including exploring approaches such as individual counselling and the benefits of support groups in providing affiliation and emotion support through a sense of belonging, instrumental support in the form of a safe place for discussion, practical aid such as informational support and normalisation, may be beneficial.

## Data Availability

Data will be provided upon request to the corresponding author.

## ACKNOWLEDGEMENTS

CS is the recipient of a National Health and Medical Research Council (NHMRC) Practitioner Fellowship (#1059156). JI is the recipient of an NHMRC Career Development Fellowship (#1162929). This study is funded in part by an NHMRC Project Grant (#1059515) and National Heart Foundation Future Leader Fellowship (#100833).

